# External validation of a paediatric SMART triage model for use in resource limited facilities

**DOI:** 10.1101/2023.06.05.23291007

**Authors:** Joyce Kigo, Stephen Kamau, Alishah Mawji, Paul Mwaniki, Dustin Dunsmuir, Yashodani Pillay, Cherri Zhang, Katija Pallot, Morris Ogero, David Kimutai, Mary Ouma, Ismael Mohamed, Mary Chege, Lydia Thuranira, Niranjan Kissoon, J. Mark Ansermino, Samuel Akech

## Abstract

**Introduction:** Models for digital triage of sick children at emergency departments of hospitals in resource poor settings have been developed. However, prior to their adoption, external validation should be performed to ensure their generalizability.

**Methods:** We externally validated a previously published nine-predictor paediatric triage model (SMART Triage) developed in Uganda using data from two hospitals in Kenya. Both discrimination and calibration were assessed, and recalibration was performed by optimizing the intercept for classifying patients into emergency, priority, or non-urgent categories based on low-risk and high-risk thresholds.

**Results:** A total of 2539 patients were eligible at Hospital 1 and 2464 at Hospital 2, and 5003 for both hospitals combined; admission rates were 8.9%, 4.5%, and 6.8%, respectively. The model showed good discrimination, with area under the receiver-operator curve (AUC) of 0.826, 0.784 and 0.821, respectively. The pre-calibrated model at a low-risk threshold of 8% achieved a sensitivity of 93% (95% confidence interval, (CI):89%-96%), 81% (CI:74%-88%), and 89% (CI:85%–92%), respectively, and at a high-risk threshold of 40%, the model achieved a specificity of 86% (CI:84%–87%), 96% (CI:95%-97%), and 91% (CI:90%-92%), respectively. Recalibration improved the graphical fit, but new risk thresholds were required to optimize sensitivity and specificity.

**Conclusion:** The Smart Triage model showed good discrimination on external validation but required recalibration to improve the graphical fit of the calibration plot. There was no change in the order of prioritization of patients following recalibration in the respective triage categories. Recalibration required new site-specific risk thresholds that may not be needed if prioritization based on rank is all that is required. The Smart Triage model shows promise for wider application for use in triage for sick children in different settings.

**Funder:** Wellcome Trust (UK)

## Introduction

The global burden of child mortality remains high in low and middle-income countries (LMICs). Despite significant progress globally, Sub-Saharan Africa continues to record mortality rates of 74 (95% confidence interval (CI), 68-86) deaths per 1000 live births, which is approximately 14 times higher than the mortality rate of children in Europe and North America (1, 2). These numbers, accounting for paediatric deaths outside the neonatal period, are largely attributed to infectious diseases including malaria, pneumonia, and diarrhoeal diseases which can be prevented or treated through simple interventions and training of healthcare workers (3).

Early recognition of critically ill children upon arrival to hospitals supports the attainment of the third Sustainable Development Goal (SDG) on ending preventable deaths of children by 2030, especially with increased availability and use of health facilities (4). Although the World Health Organization (WHO) has recommended the use of Emergency Triage and Treatment (ETAT) guidelines for the triage of sick children in resource-limited settings, its implementation in clinical practice has been challenging for many reasons including difficulties of scaling up the required training (5). Shortages of adequately trained frontline health workers in emergency areas has also been cited as impairing triage (6, 7). Without triage, children are frequently seen in order of arrival instead of according to the priority of illness.

Clinical prediction models that classify children on arrival to the hospital according to risk (that is, emergency, priority or non-urgent cases), can help frontline health workers identify critically ill children and prioritize care to reduce mortality and morbidity in resource-limited settings (8). Several predictive triage models have been developed but few models have been validated externally (8, 9).

Prediction models tend to perform well during internal validation, but this may not be replicated externally, therefore, it is recommended that model performance be examined in a different context (10–12). Ideally, clinical prediction models should be reproducible and generalizable to diverse patient groups and geographical locations and models that are not generalizable are a waste of limited research resources.

In this paper, we externally validate a previously developed nine-predictor pediatric triage model, referred to as the Smart Triage Model (13), used for the triaging of severely ill children. The original model was developed using data from Jinja, Uganda and this study performs external validation using data from two hospitals in Kenya. We also present results of updating the prediction model using recalibration-in-the-large, a method which adjusts the average predicted probability to be equal to the observed event rate.

## Methods

Model external validation adhered to Transparent Reporting of a multivariable prediction model for Individual Prognosis or Diagnosis (TRIPOD) guidelines on developing, validating, or updating a multivariable clinical prediction model (14) (Supplementary file S1).

### Study registration

The study was registered in Clinical Trials.gov, identifier: NCT04304235, on 11^th^ March 2020.

### Study population and design

The Smart Triage model was developed in a study conducted at the pediatric emergency department (ED) in Jinja Regional Referral Hospital (JRRH), a public hospital within the Uganda Ministry of Health, between April 2020 and March 2021 (9). The Smart Triage model equation is a multiple logistic regression model that includes nine predictor variables which were selected using bootstrap stepwise regression and clinical expertise. The model can be used for rapid identification of critically ill children at triage and can be integrated into any digital platform.

The validation of the Smart Triage model was performed using a dataset from two sites in Kenya, the Mbagathi County Hospital (Hospital 1) and Kiambu County Referral Hospital (Hospital 2), independently and in combination. Both hospitals are located in resource-limited urban settings and the pediatric outpatient departments (OPD) in each of the two hospitals receive approximately 20,000 patients per year with an admission rate of 10% and 7% respectively.

The study was approved by institutional review boards at Kenya Medical Research Institute Scientific Ethics Review Unit KEMRI (SERU#3958) in Kenya and the University of British Columbia in Canada (ID: H19-02398; H20-00484).

### Sampling and Eligibility

Children seeking medical care at the OPD between 8:00 am and 5:00 pm on weekdays were screened for eligibility at JRRH, Uganda, and the two Kenya hospitals. In Kenya, assent was required for children above 13 years of age in addition to caregiver consent. Both sites only enrolled children presenting with medical illness and written informed consent was provided by a parent or guardian prior to enrollment. Children presenting for elective surgical procedures, scheduled clinic appointments, or those coming to be reviewed at the hospital for treatment of chronic illnesses were not eligible for enrollment. Emergency cases who were eligible but required immediate emergency treatment (within 15 minutes of arrival) were not enrolled to the study since care of the participant was prioritized. At both Kenya hospitals, three study nurses and two timekeepers were recruited and trained to conduct study-specific procedures. The timekeepers recorded arrival time for all patients arriving with an acute illness to the OPD and used a systematic sampling method based on 30-minute time cut-offs to determine the next participant to approach for recruitment to the study. The study nurses verified patient eligibility, and if the first participant in a given cut off was not eligible or did not consent, they would examine the next participant in the same cut off depending on order of arrival. Study information was collected from those eligible after obtaining informed consent (6).

### Data collection and management

The data collection method used to develop the initial model was repeated in hospitals 1 and 2(6). Data was collected using a custom-built Android mobile application installed on a password secured tablet. A Masimo iSpO2® Pulse Oximeter was connected to the tablet to measure the pulse oximetry and heart rate. Data on the tablet was stored in an encrypted format. Each day, data collected using the tablets was uploaded to a REDCap (Research Electronic Data Capture)(15) database in a central server housed at KEMRI Wellcome Trust Research Programme (KWTRP) office, Nairobi. Standard operating procedures were developed and used for data collection and these are available on the <u>Paediatric Sepsis CoLab</u> <u>Dataverse</u> (16).

### Primary outcome

The primary outcome was defined as a composite of any one or more of the following: hospital admission for more than 24 hours as determined independently by the hospital clinician on duty (who was not part of the study team), mortality within 24 hours of admission or readmission, or referral within 48 hours to any other hospital after being seen by a hospital clinician/s, determined through follow up calls made 7 days after the initial hospital visit.

### Predictors

The nine predictors included in the JRRH final model equation were square root of age in months, heart rate, temperature, mid-upper arm circumference, transformed oxygen saturation (using the concept of virtual shunt(17), parental concern, difficulty breathing, oedema, and pallor. The model was developed by multivariable logistic regression and the model equation which was referred to as the Smart Triage model equation is: logit (p) = -32·888 + (0·252, square root of age) + (0·016, heart rate) + (0·819, temperature) + (-0·022, mid-upper arm circumference) + (0·048 transformed oxygen saturation) + (1·793, parent concern) + (1·012, difficulty breathing) + (1·814, oedema) + (1·506, pallor) (13).

### Sample size

We used a four-step procedure implemented in the pmsampsize R package (18) to determine the minimum required sample to perform validation of the model. Assuming an input C-statistic of 0.8, a prevalence of admission of 0.05, a Cox-Snell R-squared of 0.0697 based on 0.05 acceptable difference in apparent and adjusted R-squared, 0.05 margin of error in estimation of intercept, and a minimum number of events per predictor parameter(EPP) of 7, the minimal sample size required was 1117 participants with 64 events (19–22).

Observations with missing outcomes or missing 25% of the predictor variables were excluded from this analysis. All other missing values were imputed using K-Nearest neighbors (23).

### Model validation and calibration

The Smart Triage model equation was applied to data from the two Kenya hospitals separately and then to the two data sets combined. The discriminative ability of the model was assessed using the area under the receiver-operator curve (AUC) and interpretation was based on the following criteria: non-informative (AUC ≤ 0·5), poor discrimination (0.5 < AUC < 0·7), and good discrimination (AUC ≥ 0.7) (24). Calibration, which is a measure of agreement between observed and predicted values, was assessed graphically by estimating the slope on a calibration plot of predicted versus observed outcome rates in each decile of predicted probability. A calibration slope of 1 and intercept of 0 is considered ideal (25), a slope value greater than 1 shows that the predictions are too narrow to distinguish a positive and negative outcome (26). We interpreted the calibration slope using the following criteria: non-informative (slope ≤ 0·5), poor calibration (0.5 < slope < 0·7), and good calibration (slope ≥ 0·7). We assessed calibration using the Hosmer-Lemeshow statistic (27) and a p-value <0.05 was considered significant.

To improve the calibration plot of predicted against observed values we performed recalibration-in-the-large (re-estimation of the model intercept) (28). Recalibration-in-the-large adjusts the intercept of the original model on a correction factor in equation 1.

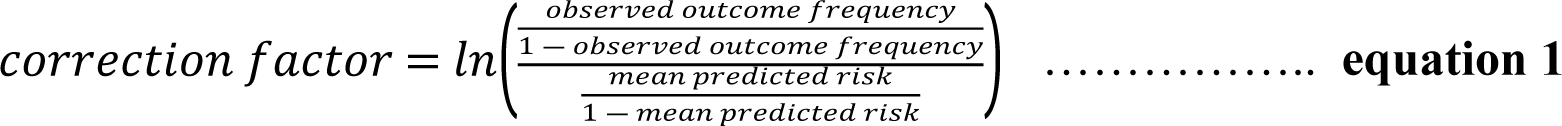

### Risk stratification

A risk classification table was used to examine the accuracy of the recalibrated model in classifying patients into triage categories. The low-risk threshold was selected to maximize sensitivity to limit misclassification of emergency and priority cases as non-urgent (avoiding false negatives) while the high-risk threshold was selected to maximize specificity to limit misclassification of non-urgent or priority cases as emergency cases (avoiding false positives) (8). We computed the true and false positive rates, negative predictive values (NPVs), and positive predictive values (PPVs) for each triage group.

## Results

### Demographic characteristics

At Hospital 1, 2680 patients were screened for eligibility between 24^th^ February 2021 and 6^th^ November 2022; 2539 patients (94.7%) met the inclusion criteria and were included in analysis. Of those that were analyzed, 226 (8.9%) had a positive primary outcome and 79.8% of all participants were aged 5 years or younger. No participants had 25% of the predictor variables missing, but 13 participants were missing the admission outcome and were thus excluded from analysis (Fig 1). The most common reason for admission was pneumonia, diagnosed using clinical signs criteria, which accounted for 55.8% of the admissions; 56.2% of all admission were male (Table 1).

**Figure 1:**
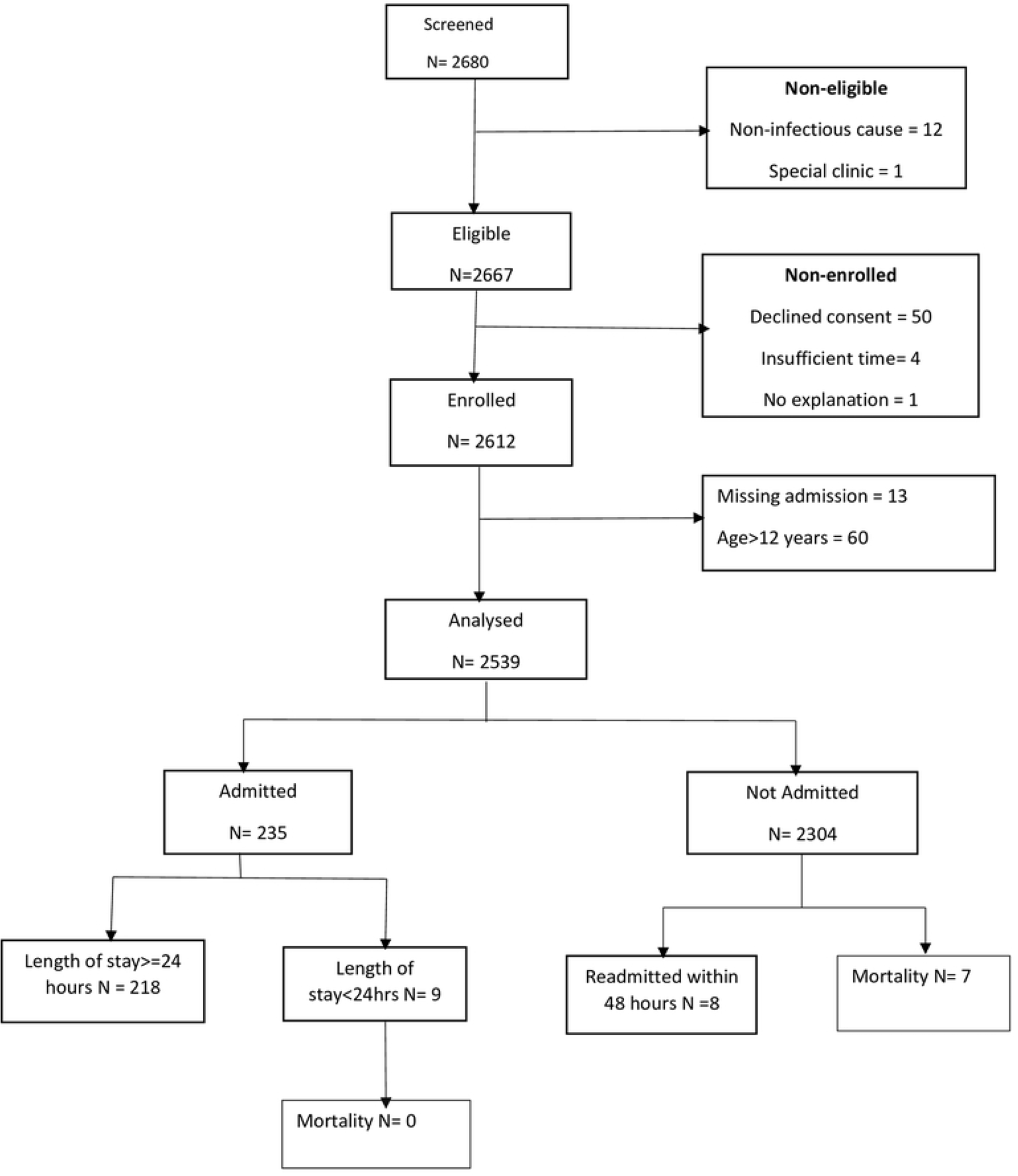
Sample population flow chart for dataset used in analysis at Mbagathi County Hospital (Hospital 1).

**Table 1:**
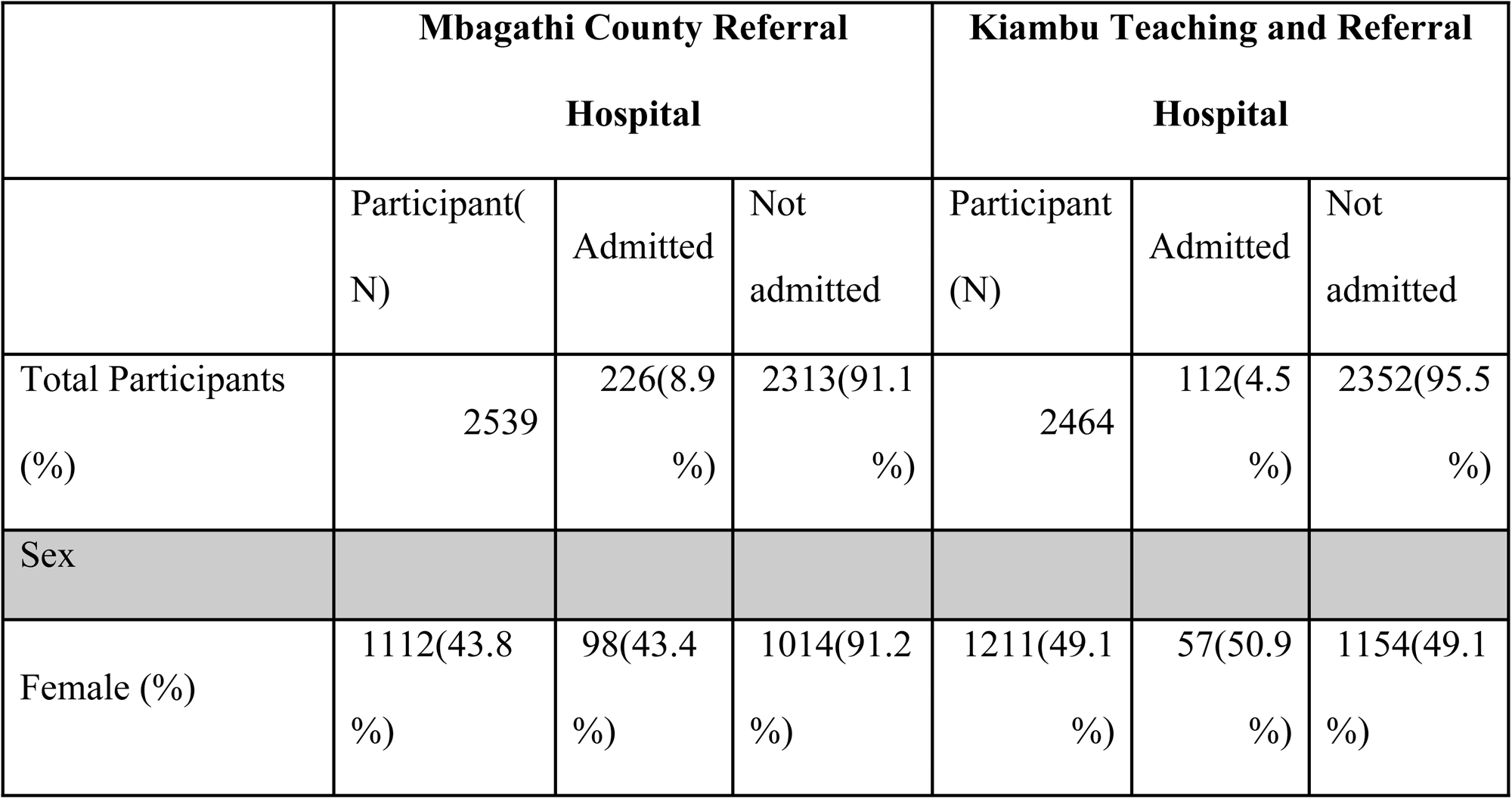
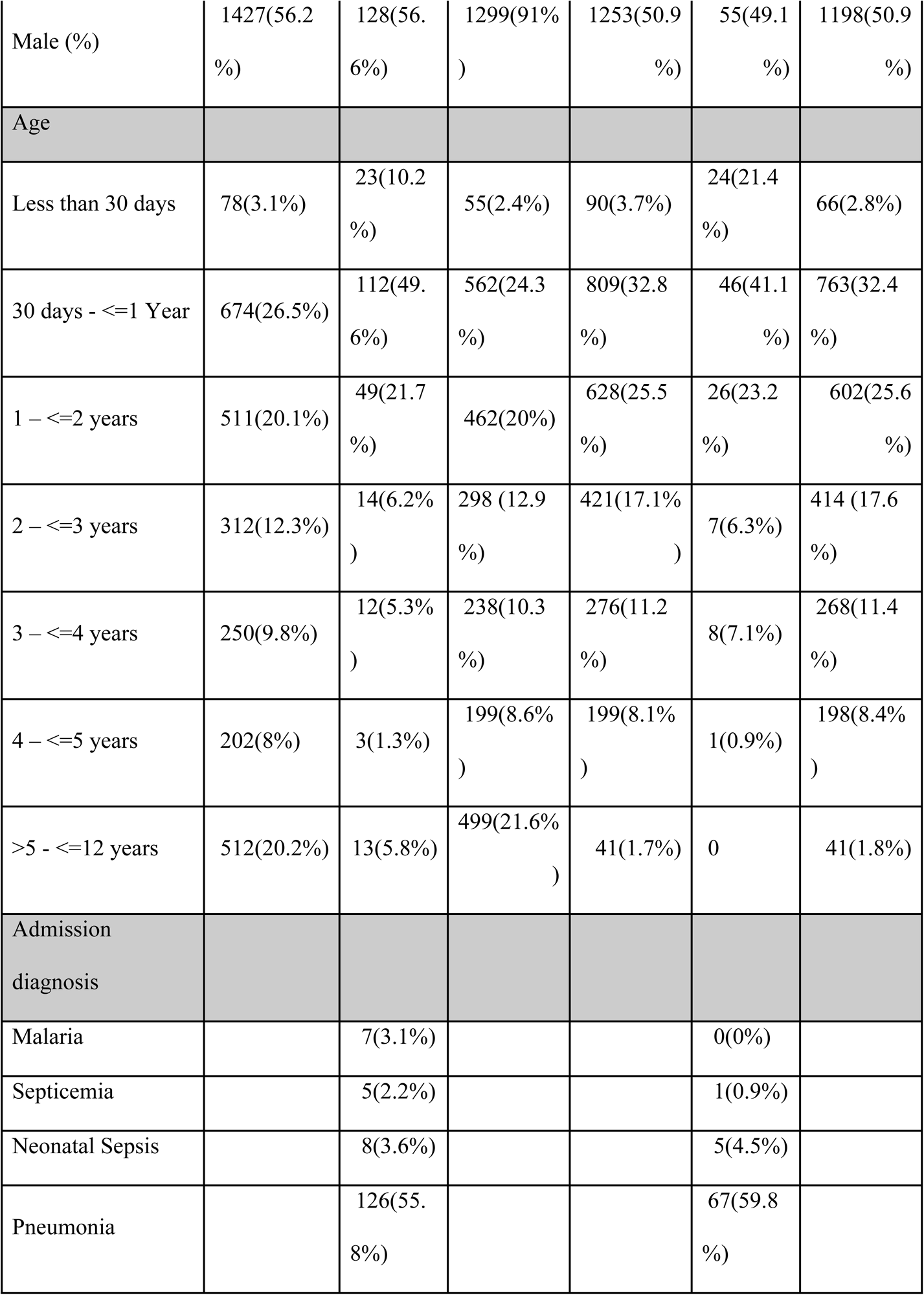
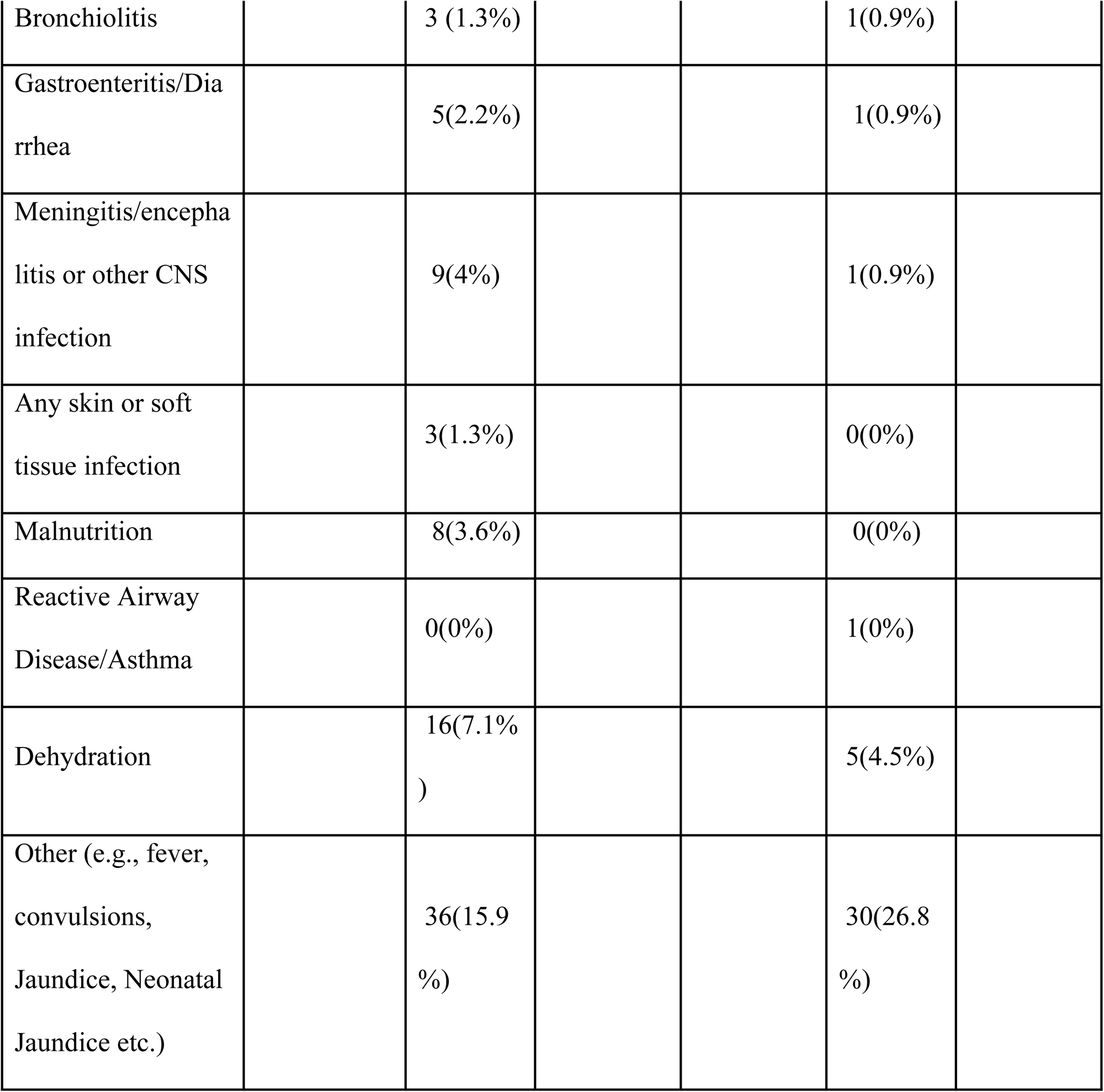
Patient’s characteristics stratified across those with and without outcome(admission)

At Hospital 2, 2671 participants were screened for eligibility between 24^th^ February 2021 and 6^th^ November 2022, of whom 2464 (92.3%) participants met the inclusion criteria and 112(4.5%) participants had a positive primary outcome. One participant had missing admission outcome and 3 withdrew from the study after enrolment and thus were excluded from this analysis. No patient had more than 25% of predictor variables missing (Fig 2).

**Figure 2:**
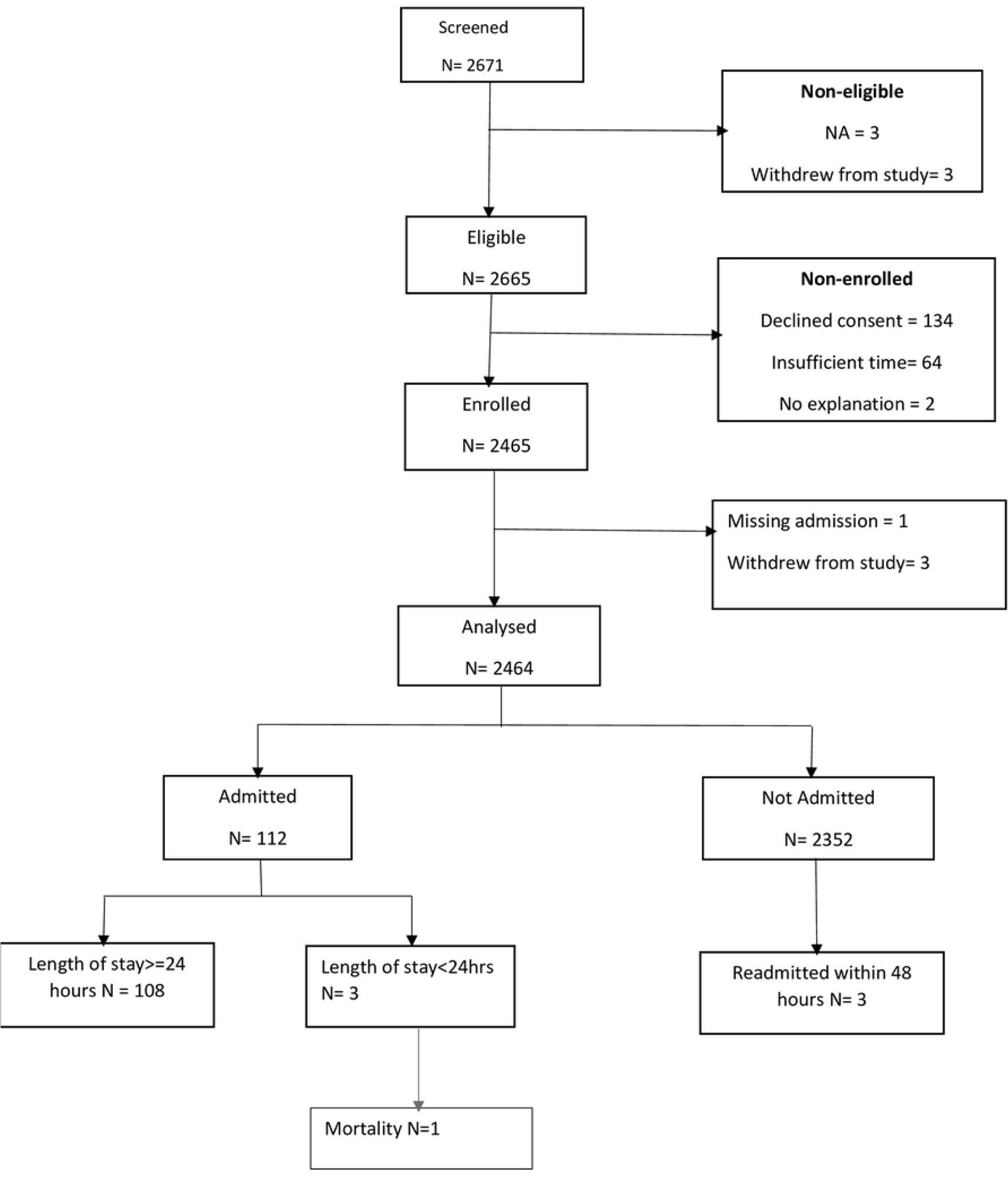
Sample population flow chart for dataset used for analysis at Kiambu County Referral Hospital (Hospital 2).

Among the 112 admitted participants, 50.9% were female and 98.3% of all the participants were aged 5 years or younger. The most common reason for admission was pneumonia which accounted for 59.8% of the admissions (Table 1). For both hospitals combined 5003 participants were analyzed and 338(6.8%) participants had positive primary outcome.

### Model performance

At Hospital 1 the model achieved good discrimination with an AUC value of 0.826 (Fig3A) and a calibration slope value of 0.72. Graphically the calibration plot improved on performing recalibration-in-the-large (Fig 4A, 4B) when the model intercept was adjusted from -32·888 to -34. 393. The Hosmer-Lemeshow test pre-calibration had a p-value < 0.00001 and on recalibration the p-value was <0.01, both p-values were below 0.05 significance level.

At Hospital 2 the model achieved good discrimination with an AUC value of 0.784 (Fig 3B) and a calibration slope value of 1.00. Graphically the calibration plot improved (Fig 4C,4D) when the new model intercept was adjusted from -32·888 to-34.201. The Hosmer-Lemeshow statistic pre-calibration had (p-value < 0.00001) and on recalibration had p-value =0.31.

**Figure 3:**
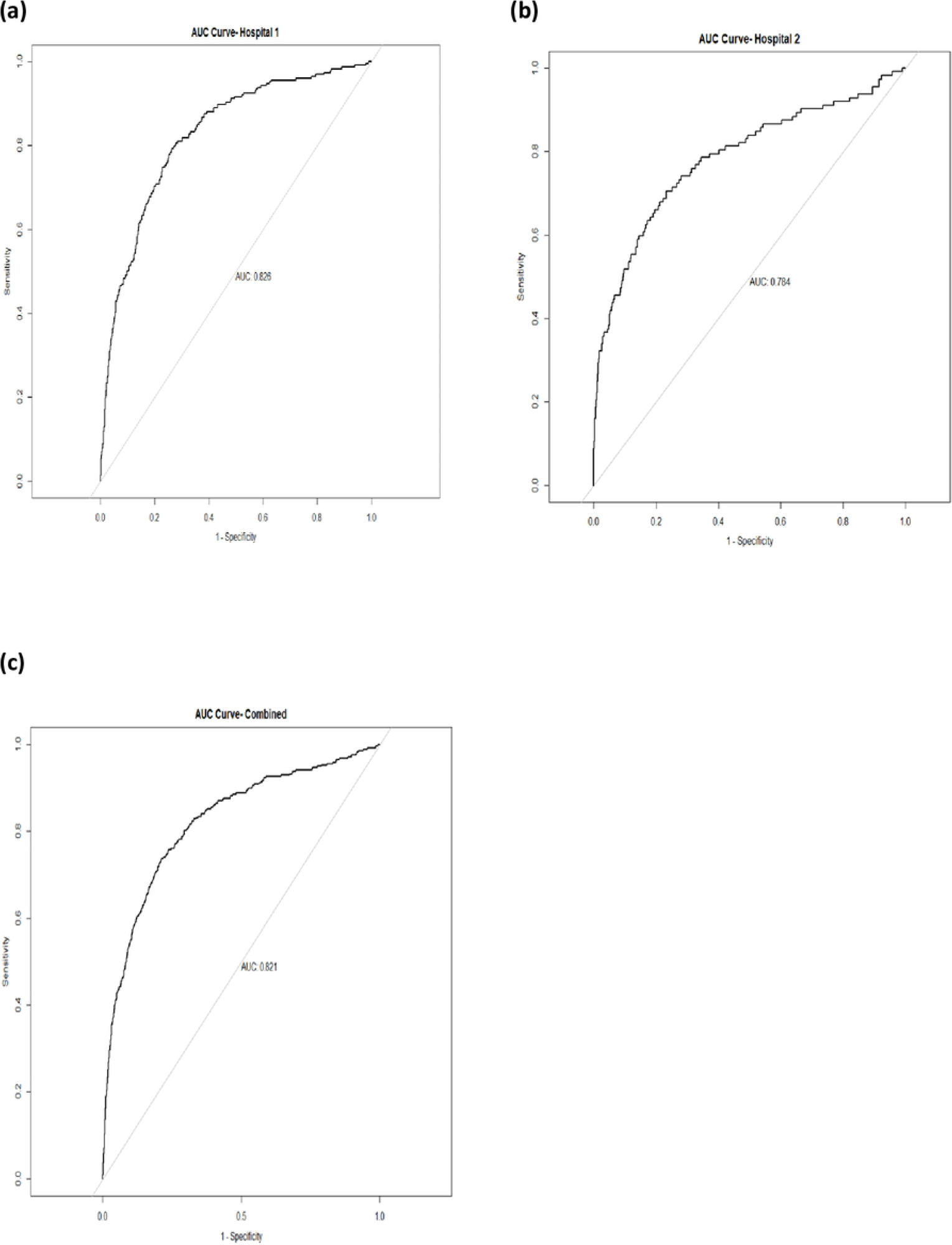
Area Under the receiver-operator curve-AUC; This shows the discrimination ability of model (C-statistic) in respective hospitals.

**Figure 4:**
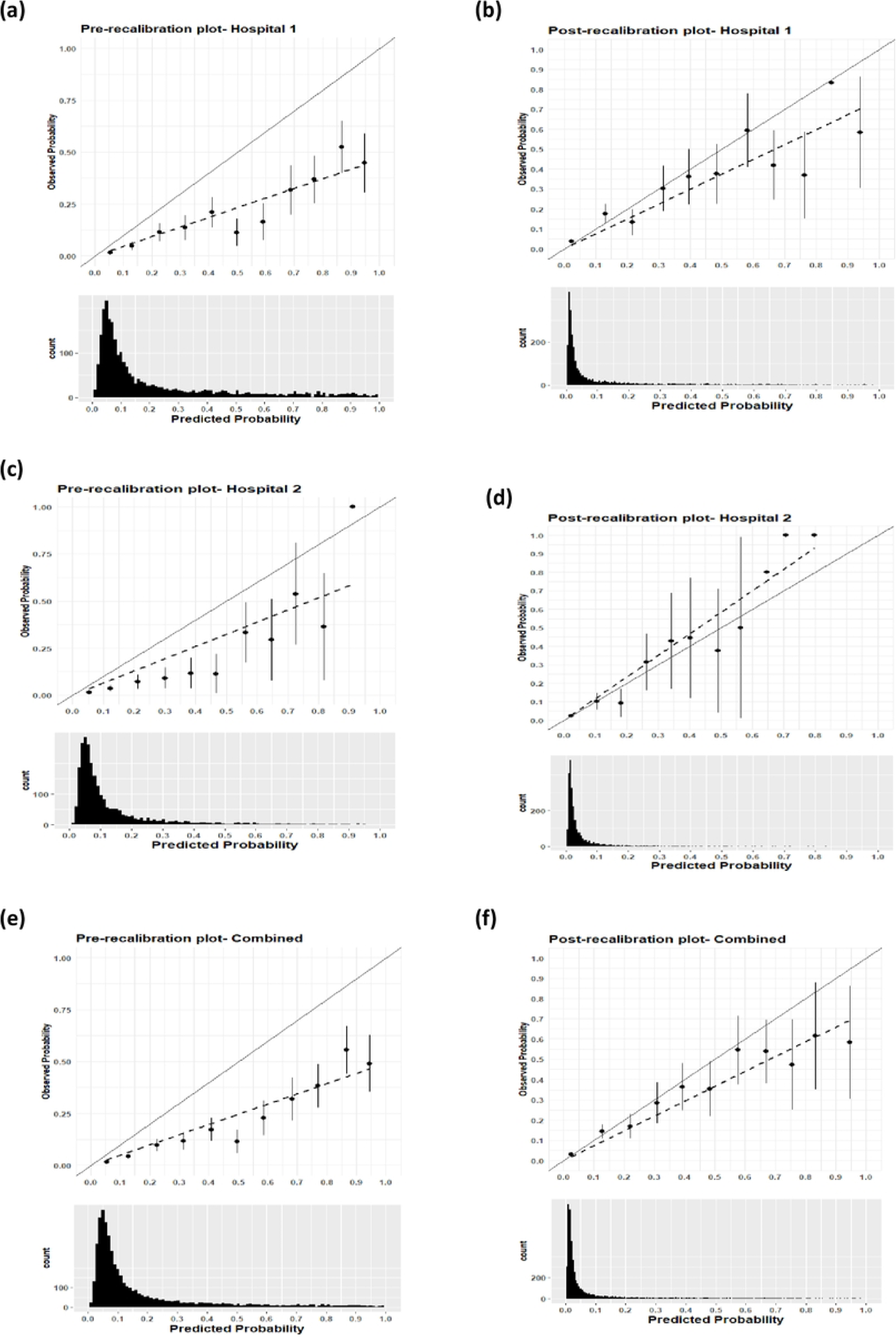
Pre-calibration and post-calibration plot; On the left are the pre-calibration plots for respective sites and on the right are post-recalibration plots for respective site; The shift of the predicted values can be explained by the histograms

For both sites combined the model achieved good discrimination with an AUC value of 0.821(Fig 3C) and calibration slope value of 0.796. On performing recalibration, the model calibration slope improved (Fig 4E, 4F) when the new model intercept was adjusted from - 32·888 to -34.325. The Hosmer-Lemeshow test pre-calibration had a (p-value < 0.00001) and on recalibration had p-value <0.05.

### Risk classification

At Hospital 1 before performing recalibration the low-risk threshold of 0.08 achieved a sensitivity (CI) of 93% (89% to 96%) and at a higher risk threshold of 0.4 the model achieved specificity (CI) of 86% (84% to 87%). On performing recalibration, the predicted values shifted toward zero and this required a new set of thresholds to maintain the numbers assigned to the various triage categories; at a low-risk threshold of 0.026 the model achieved sensitivity of 90% CI (86% to 93%) and at high-risk threshold of 0.13 the model achieved specificity of 86% CI (85% to 88%). Before calibration 60% of the admitted participants were categorized in the emergency category and this was unchanged with the use of the optimized thresholds (Table 2).

**Table 2:** Pre-calibration and post-recalibration risk classification table at selected thresholds with low-risk threshold selected to maximize sensitivity (limit misclassification of emergency and priority cases as non-urgent (avoiding false negatives)) while the high-risk threshold selected to maximize specificity (increase correct classification of emergency cases (avoiding false positives)).

| Precalibration table |  |  |  |  |  |  |  |
| --- | --- | --- | --- | --- | --- | --- | --- |
|  | Triage Category(Risk Threshold range ) | Participant n (%) | Participant with outcome, n (%) | Sensitivity(95%CI) | Specificity(95%CI) | NPV | PPV |
| Hospital 1 | Emergency(>=0.4) | 463(18%) | 136(60%) | 0.60(0.54,0.66) | 0.86(0.85,0.87) | 0.96(0.95,0.96) | 0.29(0.27,0.32) |
|  | Priority(>0.08<0.4) | 1027(40%) | 73(32)% | 0.75(0.69,0.81) | 0.77(0.75,0.79) | 0.97(0.96,0.98) | 0.24(0.22,0.26) |
|  | Non-urgent (<=0.08) | 1049(41%) | 17(8%) | 0.93(0.89,0.96) | 0.45(0.43,0.47) | 0.98(0.98,0.99) | 0.14(0.13,0.15) |
| Hospital 2 | Emergency(>=0.4) | 135(5%) | 41(37%) | 0.37(0.28,0.46) | 0.96(0.95,0.97) | 0.97(0.97,0.97) | 0.30(0.24,0.37) |
|  | Priority(>0.08<0.4) | 992(40%) | 50(45%) | 0.64(0.55,0.73) | 0.81(0.80,0.83) | 0.98(0.97,0.99) | 0.14(0.12,0.16) |
|  | Non-urgent (<=0.08) | 1337(54) | 21(19%) | 0.81(0.74,0.88) | 0.56(0.54,0.58) | 0.98(0.98,0.99) | 0.08(0.07,0.09) |
| Combined | Emergency(>=0.4) | 598(12%) | 177(52%) | 0.52(0.47,0.58) | 0.91(0.90,0.92) | 0.96(0.96,0.97) | 0.23(0.27,0.33) |
|  | Priority(>0.08<0.4) | 2019(40%) | 123(36%) | 0.67(0.62,0.72) | 0.83(0.82,0.84) | 0.97(0.97,0.98) | 0.23(0.21,0.24) |
|  | Non-urgent (<=0.08) | 2386(48%) | 38(11%) | 0.89(0.85,0.92) | 0.50(0.49,0.52) | 0.98(0.98,0.99) | 0.12(0.11,0.12) |
| Recalibration table |  |  |  |  |  |  |  |
| Hospital 1 | Emergency (>=0.130) | 458(18%) | 135(60%) | 0.60(0.53,0.66) | 0.86(0.85,0.87) | 0.96(0.95,0.96) | 0.30(0.27,0.33) |
|  | Priority(>0.026<0.130) | 750(30%) | 68(30%) | 0.71(0.65,0.77) | 0.79(0.78,0.81) | 0.97(0.96,0.97) | 0.25(0.23,0.27) |
|  | Non-urgent(<=0.026) | 1331(52%) | 23(10%) | 0.90(0.86,0.94) | 0.57(0.55,0.59) | 0.98(0.98,0.99) | 0.17(0.16,0.18) |
| Hospital 2 | Emergency (>=0.108) | 210(9%) | 51(46%) | 0.46(0.36,0.55) | 0.93(0.92,0.94) | 0.97(0.97,0.98) | 0.24(0.20,0.29) |
|  | Priority(>0.020<0.108) | 1088(44%) | 43(38%) | 0.57(0.48,0.66) | 0.86(0.85,0.88) | 0.98(0.97,0.98) | 0.14(0.17,0.19) |
|  | Non-urgent(<=0.020) | 1166(47%) | 18(16%) | 0.84(0.77,0.9) | 0.49(0.47,0.51) | 0.99(0.98,0.99) | 0.07(0.06,0.08) |
| Combined | Emergency (>=0.106) | 744(15%) | 199(59%) | 0.59(0.54,0.64) | 0.88(0.87,0.89) | 0.97(0.96,0.97) | 0.27(0.25,0.29) |
|  | Priority(<0.020<0.106) | 1904(38%) | 101(30%) | 0.69(0.65,0.74) | 0.82(0.80,0.83) | 0.97(0.97,0.98) | 0.21(0.20,0.23) |
|  | Non-urgent(<=0.020) | 2355(47%) | 38(11%) | 0.89(0.85,0.92) | 0.50(0.48,0.51) | 0.98(0.98,0.99) | 0.11(0.11,0.12) |

At Hospital 2 before performing recalibration, the model achieved sensitivity of 81 CI (74% to 88%) and specificity of 96% CI (95% to 97%). On performing recalibration with a low-risk threshold of 0.020 the model achieved sensitivity of 84% CI (77% to 90%) and at a high-risk threshold of 0.108 the model achieved specificity of 93% CI (92% to 94%). Before calibration, 37% of the admitted participants were categorized in the emergency category and on recalibration, 46% were categorized in the emergency category (Table 2).

For both hospitals combined before performing recalibration at low-risk threshold of 0.08 the model achieved a sensitivity of 89% CI (85% to 92%) and at the higher risk threshold of 0.4 the model achieved a specificity of 91.0% CI (90% to 92%). On performing recalibration at low-risk threshold of 0.020 the model achieved sensitivity of 89% CI (85% to 92%) and at high-risk threshold of 0.106 the model achieved a specificity of 88% (87% to 89%). Before calibration 53% of the admitted participants were categorized in the emergency category and on recalibration, 59% were categorized as emergency (Table 2).

## Discussion

### Key results

Performance of the Smart Triage model showed stable discrimination in all the three sets of data which suggests that for a pair of randomly selected children, the model would assign the higher risk score to the one with positive outcome compared to an individual with a negative outcome. On graphically assessing calibration of the predicted against observed outcomes, the graphical plot deteriorated in all the three sets of data shifting towards overprediction.

Recalibration-in-the-large improved the calibration plot, but this required a change in risk thresholds to optimize sensitivity and specificity and organize patients into clinically manageable risk classification triage categories.

The stability of discrimination and increasing overprediction or underprediction has been observed in parallel prediction validation studies (29–32). The change in performance often occurs because of data variation in the patient population including changes in the outcome rate, disease incidence, and prevalence across different regions, patient case mix and clinical practice (33–36). In our case, this was attributed to fewer admissions in both sites compared to the hospital where the model was developed and the lower number of participants with pallor (anaemia), one of the predictor variables. The lower prevalence of pallor at the study hospitals compared to the primary hospital is expected because the primary hospital was located in a malaria endemic zone (anaemia is a common complication of malaria) while the validation hospitals are in an area without local malaria transmission. When an algorithm is developed in a setting with high disease prevalence it may systematically overestimate risk when used in settings with lower disease incidence (37).

Appropriately implemented prediction models can be helpful in supporting decision making to improve patient outcomes and prioritize allocation of resources. Integrating predictive analytics into electronic health record systems enables the use of predictive models and can allow incorporation of probability-based tools into clinical decision support systems (29, 33, 38). In this study we performed external validation on an existing paediatric triage model (Smart Triage Model) which could be integrated into a health record systems in LMIC, after appropriate contextualization, to strengthen the triage system.

While recalibration improved the fit of the model, it also required an adjustment of risk thresholds. This would limit more generalized adoption of the model if recalibration was required at each site. Future work will investigate options to predict optimal calibration based on easily collected site specific information (such as admission rate or malaria prevalence) or the option of using the same model and thresholds, even if calibration is not optimal.

### Limitations

A significant limitation of this study was the choice of hospital admission as primary outcome measure as it is not the most robust measure of illness severity. However, to exclude admitted cases that lacked severe illness we included only children admitted more than 24 hours. To capture children who were sent home but had severe illness we included children who were readmitted within 48 hours determined from a follow up call made 7 days after discharge. We also included mortality within 24 hours of admission or readmission to still capture the severely ill.

Patient characteristics tend to change over time and in different geographical settings. There may be arguments that the findings need replication in more geographical settings with varied event rate, case mix, and hospital contexts but we believe that findings of external validation of the Smart Triage model from these two hospitals is reassuring on its adaptability to varied contexts and patient populations.

## Conclusion

The Smart Triage model showed good discrimination on external validation but required modest recalibration to improve the graphical fit of the calibration plot. On recalibration, new site-specific set of thresholds were required to maintain the same sensitivity and specificity across the triage categories. There was no significant change in the distribution of patients into the three triage categories, which alleviates concerns about model updating for prediction if prioritization based on rank is all that is required. Future research could examine whether the Smart Triage model can be applied to different populations if only the risk thresholds are adjusted without recalibration. The Smart Triage model shows promise for wider application for use in triage for sick children in different settings.

## Declarations

### Author contributions

SA, AM, NK DD, and JMA designed the study. SK, JK, YP, KP DD, DK, MO, IM, MC, LT SA and JMA were responsible for study implementation and monitoring. DD was responsible for software development. JK, DD, SK, MO and PM were responsible for data management. JK, CZ conducted the statistical analysis. JK wrote the original manuscript, which was critically reviewed and revised by the team. All authors approved the final manuscript.

### Data sharing

The data studied and data dictionary is available upon request and users who wish to reuse the source data can make a request through KEMRI-Wellcome Trust Research Program data governance committee. This committee can be contacted at.

## Data Availability

https://doi.org/10.7910/DVN/942AND

## Acknowledgements

We would like to thank the administration in both Mbagathi County Hospital and Kiambu County Referral Hospital and all participants and caregivers for their contribution in making this study a success. We would like to thank the research team in both hospitals i.e., Sidney Korir, Kevin Bosek, Verah Karasi, John Mboya, Brian Ochieng, Angela Wanjiru, Mercy Mutuku, Anastasia Gathigia, Esther Muthoni, Emmah Kinyanjui, Felix Kimani, Faith Wairimu for their hard work and dedication in data collection. This research was supported by Wellcome Trust.

